# SARS-CoV-2 B.1.1.529 variant (Omicron) evades neutralization by sera from vaccinated and convalescent individuals

**DOI:** 10.1101/2021.12.08.21267491

**Authors:** Annika Rössler, Lydia Riepler, David Bante, Dorothee von Laer, Janine Kimpel

## Abstract

Recently, the severe acute respiratory syndrome coronavirus 2 (SARS-CoV-2) variant B.1.1.529 (Omicron) has been described.

Here, we analyze titers of neutralizing antibodies of sera from convalescent or vaccinated individuals against the new B.1.1.529 variant and compared them with titers against other Variants of Concern (B.1.1.7, B.1.351, B.1617.2) using replication competent SARS-CoV-2 variants.

We found that sera from vaccinated individuals neutralized the B.1.1.529 variant to a much lesser extent than any other variant analyzed. Neutralization capacity against B.1.1.529 was maintained best against sera from super immune individuals (infected and vaccinated or vaccinated and infected).

---

During the severe acute respiratory syndrome coronavirus 2 (SARS-CoV-2) pandemic several new virus variants have emerged, whereby the virus has become more contagious. However, efficient immune escape has not been observed and vaccines have remained effective. Most recently, the B.1.1.529 variant (Omicron) has been described, which the World Health Organization (WHO) has classified as Variant of Concern (VOC) on November 26^th^ 2021^1^

B.1.1.529 is characterized by a large number of mutations with 26-32 changes in the spike (S) protein.^2^ As many of these mutations are in regions known to be involved in immune escape, we studied whether sera from vaccinated or convalescent individuals can neutralize this new VOC. The observation that B.1.1.529 is more likely to cause reinfections than previous variants indeed suggests some level of immune escape.^3^

We selected sera from patients after an infection with B.1.1.7 (n=10), B.1.351 (n=8) or B.1.617.2 (n=7) or from individuals after homologous prime/boost vaccination with Spikevax (Moderna, n=10), ChAdOx1 (Vaxzevria, AstraZeneca) (n=10), or BNT162b2 (Comirnaty, Pfizer/BioNTech) (n=20) and after heterologous ChAdOx1 prime/BNT162b2 boost vaccination (n=20) (see Table S1 and S2 in Supplementary Appendix). For all sera, we determined titers of neutralizing antibodies against B.1.1.7, B.1.351, B.1.617.2, and B.1.1.529 variants using a focus forming assay with replication competent SARS-CoV-2 viruses as described previously.^4^ In addition, we used sera from super immune individuals (n=5 for infected and subsequently vaccinated, n=5 for vaccinated and subsequently infected, see Table S3 in Supplementary Appendix) and analyzed neutralizing antibody titers against B.1.617.2 and B.1.1.529.

Figure 1 shows that sera from vaccinated individuals neutralized the B.1.1.529 variant to a much lesser extent than any other variant analyzed (B.1.1.7, B.1.351, B.1.617.2). We found some B.1.1.529 cross-neutralization in individuals vaccinated with either the homologous BNT162b2 or the heterologous ChAdOx1 prime/BNT162b2 boost regimen but not after homologous ChAdOx1 vaccination. Furthermore, the sera from convalescent individuals largely failed to neutralize B.1.1.529 although cross-neutralization was observed against other variants.

**Figure 1.**
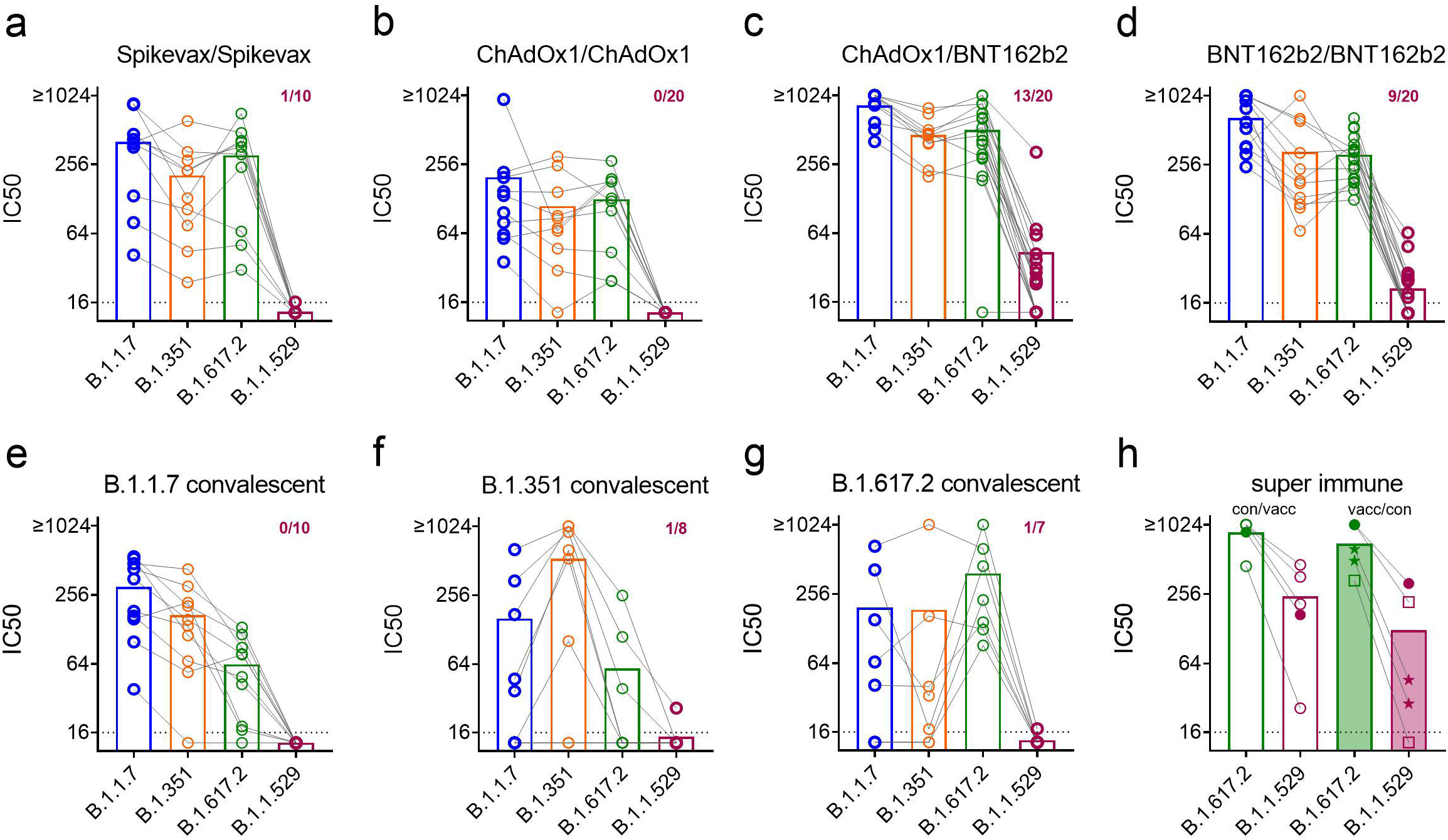
B.1.1.529 is neutralized worse compared to other Variants of concern. Sera from individuals vaccinated with two doses Spikevax (a), two doses ChAdOx1 (b), a ChAdOx1 prime followed by a BNT162b2 boost (c), two doses BNT162b2 (d), or recovered from an infection with B.1.1.7 (e), B.1.351 (f), B.1.617.2 (g) were analyzed for 50% neutralization titers (IC50) against B.1.1.7 (blue), B.1.351 (orange), B.1.617.2 (green), and B.1.1.529 (purple). Super immune sera from convalescent/vaccinated or vaccinated/convaselcent individuals (h) were analyzed for IC50 against B.1.617.2 and B.1.1.529. Dotted lines indicate detection limit. Numbers in a-g indicate proportion of positive sera for B.1.1.529 variant (>1:16). (h) ⍰ = Spikevax/Spikevax vaccinated; ★ = ChAdOx1/ChAdOx1 vaccinated; ⍰ = BNT162b2 vaccinated; ⍰ = BNT162b2/BNT162b2 vaccinated.

However, all sera from super immune individuals that had been infected and vaccinated once or twice with BNT162b2 or that had been vaccinated and subsequently were infected were able to neutralize B.1.1.529, although to a lesser degree than B.1.617.2.

Whether B.1.1.529 will become dominant and cause significant morbidity and mortality in future will depend on its transmissibility and pathogenicity. However, although a third booster vaccination with BNT162b2 may increase the level of cross-neutralizing antibodies to B.1.1.529, the presented data strongly advocate the rapid development of new, variant-adapted vaccines.

## Data Availability

All data produced in the present study are available upon reasonable request to the authors.

## Notes

### Competing Interest Statement

The authors have declared no competing interest.

### Funding Statement

This study did not receive any funding.

### Author Declarations

The ethics committee (EC) of the Medical University of Innsbruck has approved the study with EC numbers: 1330/2020, 1093/2021, 1168/202, 1191/2021, and 1197/2021.

## References

1. World Health Organization. Classification of Omicron (B.1.1.529): SARS-CoV-2 Variant of Concern. 26.11.2021; Available from: https://www.who.int/news/item/26-11-2021-classification-of-omicron-(b.1.1.529)-sars-cov-2-variant-of-concern.

2. Network for Genomic Surveillance in South Africa (NGS-SA). SARS-CoV-2 Sequencing Update 26 November 2021. Available from: https://www.nicd.ac.za/wp-content/uploads/2021/11/Update-of-SA-sequencing-data-from-GISAID-26-Nov_Final.pdf

3. Pulliamn JRC, van Schalkwyk C, Govender N et al. Increased risk of SARS-CoV-2 reinfection associated with emergence of the Omicron variant in South Africa. Dec 2, 2021. (https://doi.org/10.1101/2021.11.11.21266068) preprint.

4. Riepler L, Rössler A, Falch A et al. Comparison of 4 SARS-CoV-2 neutralization assays. Vaccines (Basel) 2020; 9(1):13.

