## Supplementary Materials for "SARS-CoV-2 B.1.1.529 variant (Omicron) evades neutralization by sera from vaccinated and convalescent individuals"

**Supplementary Appendix**

### List of Investigators

Annika Rössler

Lydia Riepler

David Bante

Dorothee von Laer

Janine Kimpel

Institute of Virology, Department of Hygiene, Microbiology and Public Health, Medical University of Innsbruck, Peter-Mayr-Str. 4b, 6020 Innsbruck, Austria

### Methods

*Ethics statement*

The ethics committee (EC) of the Medical University of Innsbruck has approved the study with EC numbers: 1330/2020, 1093/2021, 1168/202, 1191/2021, and 1197/2021.

*Focus forming neutralization assay*

Focus forming neutralization assays were performed as previously described with the following modifications.[1] Briefly, sera were heat inactivated for 30 minutes at 56°C and subsequently centrifuged for 5 minutes at 8,000 rpm in a tabletop centrifuge. Sera were four-fold diluted in duplicate samples in complete medium with 2% fetal calf serum (FCS) starting with a 1:16 dilution. Virus (B.1.1.7: isolate C69.1, GISAID ID EPI_ISL_3277382; B.1.351: isolate C24.1, GISAID ID EPI_ISL_1123262; B.1.617.2: isolate SARS-CoV-2-hCoV-19/USA/NY-MSHSPSP-PV29995/2021, GISAID ID EPI_ISL_2290769; B.1.1.529: isolate E16.1, GISAID ID EPI_ISL_6902053) was added resulting in around 100-200 infected cells in the control wells without serum and mixes were incubated for 1 hour at 37°C. Subsequently, serum/virus mixtures were transferred to confluent Vero cells stably overexpressing TMPRSS2 and ACE2. Two hours after infection, inoculum was removed and fresh medium was added. Ten hours after infection, cells were fixed for 5 minutes with 96% ethanol and subsequently stained using the serum of a convalescent patient and an Alexa Fluor Plus 488-conjugated goat anti-human IgG secondary antibody (Invitrogen, Thermo Fisher Scientific, Vienna, Austria). Plates were analyzed using an ImmunoSpot S6 Ultra-V reader and FluoroSpot software (CTL Europe GmbH, Bonn, Germany). Continuous 50% neutralization titers were calculated in GraphPad Prism 9.0.1 (GraphPad Software, Inc., La Jolla, CA, USA) using a non-linear regression. Titers >1:16 were regarded as positive.

### Table S1. Patient characteristics convalescent sera

| Infection | Vaccination | Age [years] |
| --- | --- | --- |
| B.1.1.7 | - | 60-69 |
| B.1.1.7 | - | 70-79 |
| B.1.1.7 | - | 60-69 |
| B.1.1.7 | - | 30-39 |
| B.1.1.7 | - | 50-59 |
| B.1.1.7 | - | 30-39 |
| B.1.1.7 | - | 50-59 |
| B.1.1.7 | - | 20-29 |
| B.1.1.7 | - | 40-49 |
| B.1.1.7 | - | 50-59 |
| B.1.351 | - | 80-89 |
| B.1.351 | - | 70-79 |
| B.1.351 | - | 60-69 |
| B.1.351 | - | 80-89 |
| B.1.351 | - | 80-89 |
| B.1.351 | - | 90-99 |
| B.1.351 | - | 80-89 |
| B.1.351 | - | 60-69 |
| B.1.617.2 | - | 30-39 |
| B.1.617.2 | - | 30-39 |
| B.1.617.2 | - | 40-49 |
| B.1.617.2 | - | 30-39 |
| B.1.617.2 | - | 31 |
| B.1.617.2 | - | 10-19 |
| B.1.617.2 | - | 10-19 |

### Table S2. Patient characteristics vaccinated sera

| Vaccination | Months 2nd dose | Age [years] |
| --- | --- | --- |
| ChAdOx1/ChAdOx1 | 1 | 20-29 |
| ChAdOx1/ChAdOx1 | 1 | 20-29 |
| ChAdOx1/ChAdOx1 | 1 | 20-29 |
| ChAdOx1/ChAdOx1 | 1 | 30-39 |
| ChAdOx1/ChAdOx1 | 1 | 50-59 |
| ChAdOx1/ChAdOx1 | 1 | 20-29 |
| ChAdOx1/ChAdOx1 | 1 | 50-59 |
| ChAdOx1/ChAdOx1 | 1 | 30-39 |
| ChAdOx1/ChAdOx1 | 1 | 40-49 |
| ChAdOx1/ChAdOx1 | 1 | 30-39 |
| ChAdOx1/BNT162b2 | 1 | 20-29 |
| ChAdOx1/BNT162b2 | 1 | 20-29 |
| ChAdOx1/BNT162b2 | 1 | 30-39 |
| ChAdOx1/BNT162b2 | 1 | 30-39 |
| ChAdOx1/BNT162b2 | 1 | 20-29 |
| ChAdOx1/BNT162b2 | 1 | 20-29 |
| ChAdOx1/BNT162b2 | 1 | 20-29 |
| ChAdOx1/BNT162b2 | 1 | 30-39 |
| ChAdOx1/BNT162b2 | 1 | 50-59 |
| ChAdOx1/BNT162b2 | 1 | 20-29 |
| ChAdOx1/BNT162b2 | 1 | 20-29 |
| ChAdOx1/BNT162b2 | 1 | 20-29 |
| ChAdOx1/BNT162b2 | 1 | 20-29 |
| ChAdOx1/BNT162b2 | 1 | 20-29 |
| ChAdOx1/BNT162b2 | 1 | 50-59 |
| ChAdOx1/BNT162b2 | 1 | 50-59 |
| ChAdOx1/BNT162b2 | 1 | 20-29 |
| ChAdOx1/BNT162b2 | 1 | 30-39 |
| ChAdOx1/BNT162b2 | 1 | 20-29 |
| ChAdOx1/BNT162b2 | 1 | 30-39 |
| BNT162b2/BNT162b2 | 1 | 60-69 |
| BNT162b2/BNT162b2 | 1 | 40-49 |
| BNT162b2/BNT162b2 | 1 | 40-49 |
| BNT162b2/BNT162b2 | 1 | 20-29 |
| BNT162b2/BNT162b2 | 1 | 20-29 |
| BNT162b2/BNT162b2 | 1 | 30-39 |
| BNT162b2/BNT162b2 | 1 | 20-29 |
| BNT162b2/BNT162b2 | 1 | 20-29 |
| BNT162b2/BNT162b2 | 1 | 30-39 |
| BNT162b2/BNT162b2 | 1 | 20-29 |
| BNT162b2/BNT162b2 | 1 | 30-39 |
| BNT162b2/BNT162b2 | 1 | 40-49 |
| BNT162b2/BNT162b2 | 1 | 50-59 |
| BNT162b2/BNT162b2 | 1 | 20-29 |
| BNT162b2/BNT162b2 | 1 | 20-29 |
| BNT162b2/BNT162b2 | 1 | 30-39 |
| BNT162b2/BNT162b2 | 1 | 30-39 |
| BNT162b2/BNT162b2 | 1 | 20-29 |
| BNT162b2/BNT162b2 | 1 | 20-29 |
| BNT162b2/BNT162b2 | 1 | 30-93 |
| Spikevax/Spikevax | 5 | 50-59 |
| Spikevax/Spikevax | 5 | 20-29 |
| Spikevax/Spikevax | 6 | 20-29 |
| Spikevax/Spikevax | 6 | 30-39 |
| Spikevax/Spikevax | 4 | 20-29 |
| Spikevax/Spikevax | 5 | 30-39 |
| Spikevax/Spikevax | 5 | 20-29 |
| Spikevax/Spikevax | 5 | 40-49 |
| Spikevax/Spikevax | 6 | 30-39 |
| Spikevax/Spikevax | 5 | 30-39 |

### Table S3. Patient characteristics super immune sera

| immune status* | Vaccination | Age [years] |
| --- | --- | --- |
| con/vacc | BNT162b2 | 40-49 |
| con/vacc | BNT162b2/BNT162b2 | 60-69 |
| con/vacc | BNT162b2 | 20-29 |
| con/vacc | BNT162b2 | 50-59 |
| con/vacc | BNT162b2 | 50-59 |
| vacc/con | ChAdOx1/ChAdOx1 | 50-59 |
| vacc/con | BNT162b2/BNT162b2 | 50-59 |
| vacc/con | Spikevax/Spikevax | 40-49 |
| vacc/con | Spikevax/Spikevax | 40-49 |
| vacc/con | ChAdOx1/ChAdOx1 | 40-49 |

*vacc/con: vaccination with two doses Spikevax, ChAdOx1 or BNT162b2 and subsequent infection; con/vacc: infection and subsequent vaccination either with one or two doses

### References

1.	Riepler, L., et al., *Comparison of Four SARS-CoV-2 Neutralization Assays.* Vaccines (Basel), 2020. **9**(1).
